# Longitudinal variability in mortality predicts Covid-19 deaths

**DOI:** 10.1101/2020.12.25.20248853

**Authors:** Jon O. Lundberg, Hugo Zeberg

## Abstract

Within Europe, death rates due to covid-19 vary greatly, with some countries being hardly hit while others to date are almost unaffected. It would be of interest to pinpoint the factors that determine a country’s susceptibility to a pandemic such as covid-19.

Here we present data demonstrating that mortality due to covid-19 in a given country could have been largely predicted even before the pandemic hit Europe, simply by looking at longitudinal variability of all-cause mortality rates in the years preceding the current outbreak. The variability in death rates during the influenza seasons of 2015-2019 correlate to excess mortality caused by covid-19 in 2020 (R^2^=0.48, p<0.0001). In contrast, we found no correlation between such excess mortality and age, population density, degree of urbanization, latitude, GNP, governmental health spendings or rates of influenza vaccinations.

These data may be of some relevance when discussing the effectiveness of acute measures in order to limit the spread of the disease and ultimately deaths. They suggest that in some European countries there is an intrinsic susceptibility to fatal respiratory viral disease including covid-19; a susceptibility that was evident long before the arrival of the current pandemic.

## Introduction

Seasonal fluctuations in all-cause mortality are well known with deaths typically peaking in the winter.^1^A main factor driving excess winter mortality is seasonal flu.^1^ When examining mortality across Europe we noted that some countries repeatedly exhibit excess deaths during the winter flu season while others show only minimal variations.^2^ In 2020 the peak in excess mortality was extraordinarily large, came later, and was connected with the covid-19 pandemic.^2^ Again, however, the higher death rates were unevenly spread throughout Europe. Here we decided to relate historical death rates 2015-2019 with the mortality rates seen in Europe during 2020 using a publically available database.^2^ We find a remarkable correlation between these two paramaters.

## Results and Discussion

Figure 1a shows the degree of excess all cause mortality in 25 European countries during the 10 first months of 2020. The overall excess in all cause mortality in these countries correlated well with reported deaths3 due to Covid-19 (R2 = 0.78, p <0.0001). It is clear that excess mortality varies greatly within Europe in 2020. In Fig. 1b we show that countries normally experiencing fluctuating mortality exhibited higher excess mortality also in 2020 (R2 = 0.48, p <0.0001), while those not affected during preceding years were spared. This pattern is clearly seen when longitudinal mortality rates (Z-scores) for some countries are exhibited (Fig. 1c). Thus, some countries including Spain, Belgium and Italy that were severely hit by the current pandemic also displayed high excess mortalty during the preceeding winter influenza seasons. Conversely, countries like Norway, Luxembourg and Estonia display hardly visible fluctuations in mortality rates over the entire study period including 2020.

**Figure 1.**
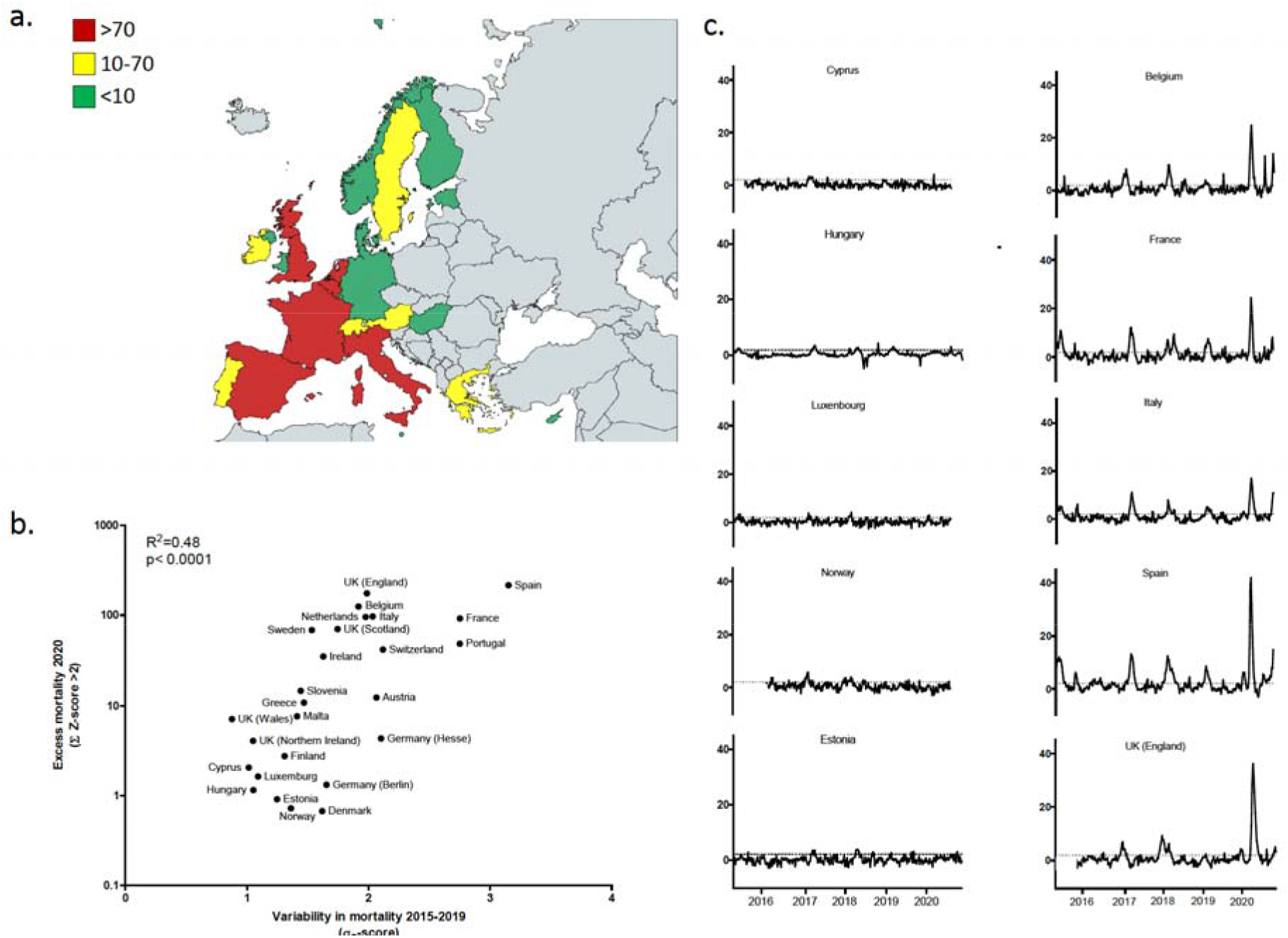
Excess mortality in Europe 2020 during the Covid-19 pandemic. (a) Map of Europe showing countries with varying degree of excess mortality during the Covid-19 outbreak in 2020, (b) variability in death rates 2015-2019 for 25 European countries plotted against the degree of excess deaths during the covid-19 outbreak and (c) longitudinal mortality patterns (Z-scores) for 10 representative countries demonstrating low (left) or high (right) death rates during 2020. Note that countries with high death rates 2020 also exhibit considerable peaks in mortality during the preceeding influenza seasons. Dotted lines represent a Z-score of 2 which here is defined as the threshold for excess mortality. Countries in gray in the map are not included in the database used. For Germany two regions (Hesse, Berlin) are included in the database and calculations. Colours in the map represent different degrees of excess mortality (sum of Z-scores >2) with red being highest, yellow intermediate and green low.

This suggests that it was in fact possible to predict high death rates caused by covid-19 even before the pandemic hit Europe simply by looking at excess mortality in a country during the preceding normal influenza seasons. Why then are some countries apparently unaffected year after year by fatal respiratory viral disease, while others suffer considerably? Possible explanations include geographical factors ^4^, population demographics, and density ^5^, genetic factors ^6^, and cultural differences along with the organization of health care and elderly nursing homes. However, we found no correlation between population age and excess mortality in 2020 and neither was there any correlation with population density, latitude, GNP, governmental health spending, degree of urbanization, or rates of influenza vaccination (Table 1 and 2). General organization and quality of health care and in particular elderly nursing homes, where a large portion of the 2020 covid-19 related deaths originated, are more complex factors that cannot be evaluated using these data alone. It will be interesting also to study if regional cultural differences across Europe might explain the pattern observed here. Again, such analysis will require more sophisticated analytical tools and datasets. In the case of the ongoing covid-19 pandemic, the degree of social interactions are influenced by governmental policies ranging from milder regulations to lockdowns. The effectiveness of these measures in preventing the spread of infection and ultimately death is currently a matter of great debate. It seems clear to date that many countries that applied very strict measures, still have experienced very high infection rates and death tolls during the current pandemic. Although the present data cannot be used to evaluate the success of governmental measures, it is noteworthy that whatever factors that drove excess mortality rates in 2020 were present already in 2015-2019, ie during a period when no measures were undertaken in any country. Thus, our data suggest that there is an intrinsic susceptibility in certain countries to excess mortality associated with respiratory viral diseases including covid-19. We suggest that knowing about such susceptibility can be of value in preparing health care systems and directing timely help to a certain region when a pandemic hits a continent.

**Table 1.**
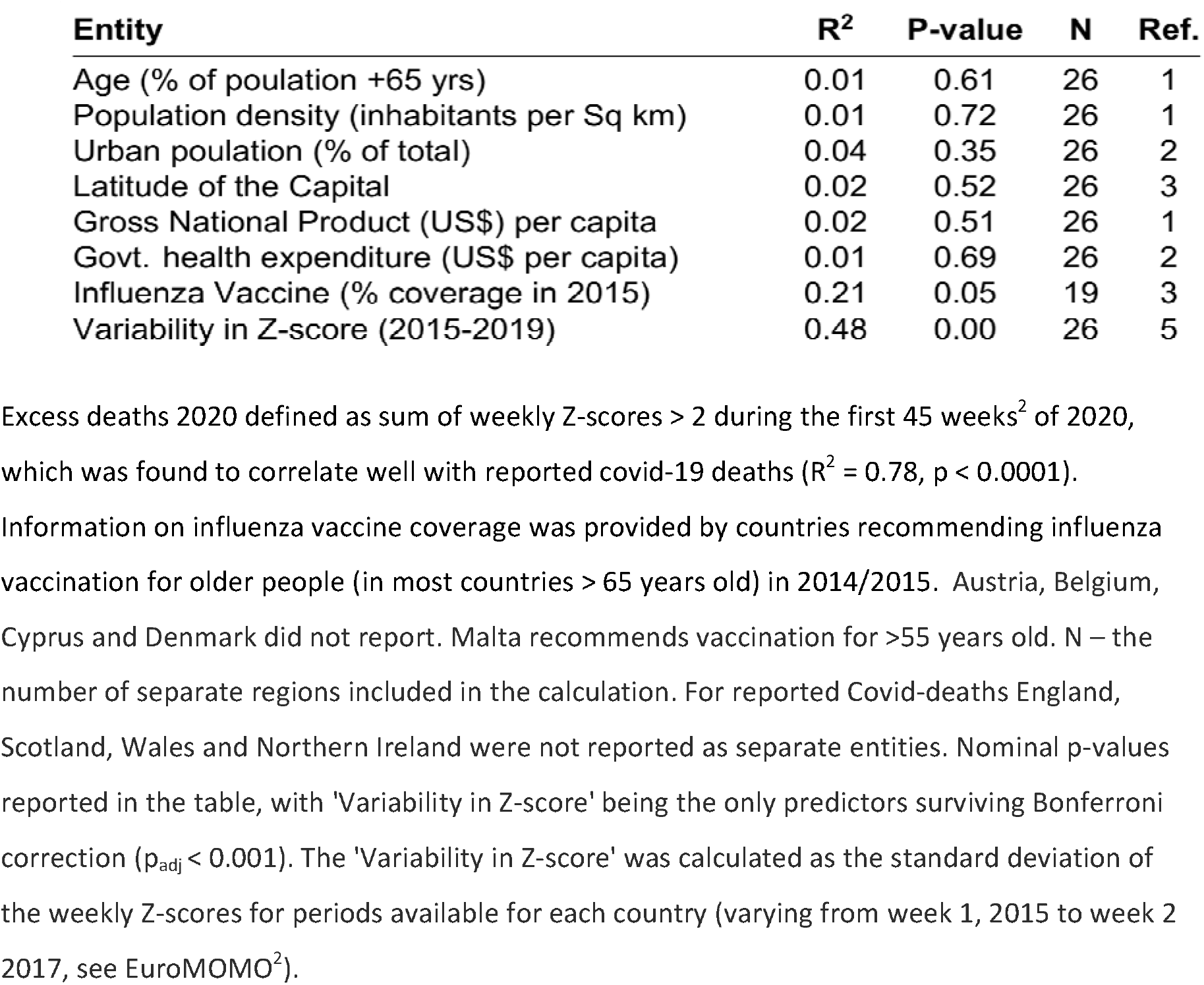
Predicitive power of some indicators for excess deaths 2020.

**Table 2.** Correlation between indicators in Table 1 and Variability in Z-score.

| **Entity** | **R<sup>2</sup>** | **P-value** | **N** | **Ref.** |
| --- | --- | --- | --- | --- |
| Age (% of poulation +65 yrs) | 0.11 | 0.09 | 26 | 1 |
| Population density (inhabitants per Sq km) | 0.00 | 0.96 | 26 | 1 |
| Urban poulation (% of total) | 0.01 | 0.58 | 26 | 2 |
| Latitude of the Capital | 0.06 | 0.22 | 26 | 3 |
| Gross National Product (US\$) per capita | 0.02 | 0.51 | 26 | 1 |
| Govt. health expenditure (US\$ per capita) | 0.00 | 0.98 | 26 | 2 |
| Influenza Vaccine (% coverage in 2015) | 0.05 | 0.37 | 19 | 4 |

## Materials and Methods

The data presented here have been extracted from the public data base European Mortality monitoring.^2^

The following sources were used for the other correlations. Population age and density (https://data.worldbank.org/indicator/SP.POP.65UP.TO.ZS); degree of urbanization, GNP and governmental health care spendings (https://www.who.int/data/gho);latitute (https://www.latlong.net); influenza vaccine rates (https://www.ecdc.europa.eu/en/seasonal-influenza/prevention-andcontrol/vaccines/vaccination-coverage).

The Map used in figure 1 was created from: https://mapchart.net/europe.html

## Data Availability

these data have been extracted from a public database www.Euromomo.eu

## Acknowledgments

The study was funded by the Swedish Research Council, Karolinska Institutet and the Swedish Heart and Lung Foundation

## References

1. Tillett HE, Smith JW, Clifford RE. Excess morbidity and mortality associated with influenza in England and Wales. Lancet 1980;1:793–5.

2. Euro Mortality Monitoring (www.euromomo.eu)

3. https://www.who.int/docs/default-source/coronaviruse/situation-reports/20201005-weekly-epi-update-8.pdf

4. Sajadi MM, Habibzadeh P, Vintzileos A, Shokouhi S, Miralles-Wilhelm F, Amoroso A. Temperature, Humidity, and Latitude Analysis to Estimate Potential Spread and Seasonality of Coronavirus Disease 2019 (COVID-19). JAMA Network Open 2020;3:e2011834.

5. Bray I, Gibson A, White J. Coronavirus disease 2019 mortality: a multivariate ecological analysis in relation to ethnicity, population density, obesity, deprivation and pollution. Public health 2020;185:261–3.

6. Ellinghaus D, Degenhardt F, et al. Genomewide Association Study of Severe Covid-19 with Respiratory Failure. N Engl J Med 2020;383:1522–34.

